# Efficacy of the wild-type/Omicron BA.1 bivalent vaccine as the second booster dose against Omicron BA.2 and BA.5

**DOI:** 10.1101/2022.11.15.22282328

**Authors:** Hitoshi Kawasuji, Yoshitomo Morinaga, Hideki Tani, Yumiko Saga, Hiroshi Yamada, Yoshihiro Yoshida, Yusuke Takegoshi, Makito Kaneda, Yushi Murai, Kou Kimoto, Akitoshi Ueno, Yuki Miyajima, Kentaro Nagaoka, Chikako Ono, Yoshiharu Matsuura, Hideki Niimi, Yoshihiro Yamamoto

**Affiliations:** Department of Clinical Infectious Diseases, Toyama University Graduate School of Medicine and Pharmaceutical Sciences, Toyama, Japan; Department of Microbiology, Toyama University Graduate School of Medicine and Pharmaceutical Sciences, Toyama, Japan; Clinical and Research Center for Infectious Diseases, Toyama University Hospital, Toyama, Japan; Department of Virology, Toyama Institute of Health, Toyama, Japan; Laboratory of Virus Control, Center for Infectious Disease Education and Research (CiDER), Osaka University, Osaka, Japan; Laboratory of Virus Control, Research Institute for Microbial Diseases (RIMD), Osaka University, Osaka, Japan; Department of Clinical Laboratory and Molecular Pathology, Toyama University Graduate School of Medicine and Pharmaceutical Sciences, Toyama, Japan

**Keywords:** bivalent, Omicron, neutralizing antibody, secondary booster, BA.1, BA.5

## Abstract

**Introduction:** In addition to the original monovalent vaccines available for SARS-CoV-2, bivalent vaccines covering wild-type (WT) and Omicron BA.1 are also available. However, there is a lack of real-world data on the effectiveness of bivalent vaccines as second boosters on the dominant Omicron sublineages, including BA.2 and BA.5.

**Methods:** This prospective longitudinal cohort study was conducted at Toyama University Hospital, a tertiary medical center in Japan. Participants (n = 565) who received the first booster vaccination were followed up until 2 weeks after the second booster dose of the monovalent mRNA-1273 (WT group, n = 168) and bivalent BNT162b2 (WT+BA.1 group, n = 23) vaccines. Participants with previous SARS-CoV-2 infections were excluded from the study. Anti-receptor-binding domain (RBD) antibody levels and neutralizing activity were measured. Vaccine-related symptoms were also assessed using a questionnaire after the second booster dose.

**Results:** The anti-RBD antibody levels after the second booster dose in the WT and WT+BA.1 group were similar (median [inter quartile], 26262.0 [16951.0–38137.0] U/mL vs. 24840.0 [14828.0–41460.0] U/mL, respectively). Although the neutralization activity of the pooled sera of the WT+BA.1 group was the lowest against BA.5, the activities against BA.2 and BA.5 were higher than those of the WT group in both pseudotyped and live virus assays. Vaccine-related symptoms, including systemic and local symptoms, were strongly correlated with anti-RBD antibody levels and neutralizing titers with significant differences.

**Conclusion:** The second booster dose of the bivalent (WT/Omicron BA.1) vaccine induced higher neutralizing activity against BA.2 and BA.5 than that of the original monovalent vaccine.

## INTRODUCTION

The emergence of the Omicron lineage of SARS-CoV-2 was first recognized in November 2021, when it spread rapidly to become globally dominant (1). Omicron (B.1.1.529 [BA.1]) and Omicron subvariants (BA.2 and BA.5), which are evolutionarily distant from the Wuhan variant (2), have large numbers of substitutions in the spike protein that can evade antibody neutralization, resulting in diminished vaccine efficacy and persistent transmission (3, 4).

Vaccination programs against COVID-19 have been conducted worldwide; however there have been inevitable variations based on the social circumstances of each country. The Japanese government recommended a voluntary fourth dose to adults older than 60 years, immunocompromised individuals, and health care workers in May 2022. A fourth dose of the original COVID-19 vaccine restores antibodies to levels observed after the third dose; however, it provides only a modest short-term boost in protection against infection (5, 6). In particular, it is extremely difficult to induce immunity against BA.5 using the original vaccines (6).

The dominant SARS-CoV-2 virus has changed constantly. Omicron BA.1 has almost been eliminated and BA.2 and BA.5 are the current dominant sublineages, whereas other sublineages such as BQ.1 and XBB are already increasing in some countries and regions (2). Bivalent vaccines are a strategy to protect against circulating variants and broaden neutralization to previous variants (7, 8). Interim data from phase 2–3 studies of bivalent vaccines covering the WT and Omicron BA.1, such as mRNA-1273.214 and Pfizer-BioNTech Bivalent, showed that higher humoral immunity against BA.2 and BA.5 was induced compared with that induced by the original monovalent mRNA-1273 vaccine, which covers only the WT (8, 9). These bivalent vaccines have already been used, but not much information is available on their clinical efficacy.

It is crucial to assess newly authorized vaccines for different communities and provide an accurate understanding of their efficacy and side effects. We conducted a prospective longitudinal study to assess the safety, immunogenicity, and reactogenicity of vaccination against SARS-CoV-2 in health care workers. In our facility, the original monovalent vaccine (mRNA-1273) or WT/Omicron BA.1 bivalent vaccine (BNT162b2) was administered as a second booster dose to health care workers. Our results revealed that the WT/BA.1 bivalent vaccine induced immunity against BA.2 and BA.5; vaccine-related symptoms were similar to those of the first booster.

## Materials and Methods

### Study design and participants

We conducted this prospective longitudinal cohort study at Toyama University Hospital, a tertiary medical center in Japan with 612 beds and 1639 healthcare workers. All participants were healthcare workers at the hospital who received the first booster (third dose) of BNT162b2 vaccine between December 6 and 23, 2021, 260 (range 248–270) days after primary BNT162b2 vaccination (first and second doses). Participants were initially invited to provide blood samples 2 weeks (2wA3D), 3 months (3mA3D), and 6 months after the third dose (6mA3D) to assess humoral responses to the third dose of vaccination, and we previously reported the safety and immunogenicity at 2wA3D of the third dose of BNT162b2 vaccine (10). In the present study, participants who provided blood samples at least one time point after the third dose were eligible for analysis and were not randomly divided into two groups: those who received the original mRNA-1273 vaccine and those who received the recently authorized Pfizer BNT162b2 bivalent Omicron BA.1-containing vaccine as the second booster (fourth dose). The former received the original mRNA-1273 vaccine on August 5, August 10, and September 2, 2022; the latter received the BNT162b2 bivalent (Wild Type [WT]/Omicron BA.1) vaccine on September 30, 2022. Eligible participants were invited to participate in this study and provided peripheral blood samples 12–16 days after the second booster vaccination (2wA4D; 2 weeks after the fourth dose). Individuals with previous SARS-CoV-2 infections were excluded from the analysis. A previous infection was determined by a documented SARS-CoV-2-positive RT-PCR result or the presence of positive anti-SARS-CoV-2 nucleocapsid antibodies. Anti-nucleocapsid antibodies in all participants were measured at 3mA3D, 6mA3D, and 2wA4D.

### Specimen collection

Serum samples were collected from the participants at 2wA3D, 3mA3D, 6mA3D, and 2wA4D. The sera were used for serological assays within 3 days of storage at 4°C or frozen at − 80°C until further verification.

### Outcome

The primary outcome measures were differences in anti-RBD antibody levels and neutralization activity against the Omicron sub-lineage (BA.2 and BA.5), between the prototype mRNA-1273 and Pfizer BNT162b2 bivalent (WT/Omicron BA.1) vaccines at 2wA4D among subjects with clarified vaccine history and known immune status before the second booster dose. Secondary outcome measures were the incidence of local and systemic adverse effects after the mRNA-1273 or BNT162b2 bivalent vaccine dose and the association of antibody levels and neutralization activity with age, sex, and adverse effects in each vaccine. In addition, the association between adverse reactions after the third and fourth doses was also evaluated because identical individuals with information about adverse reactions after the third dose could be followed in this prospective longitudinal study.

### SARS-CoV-2 pseudotyped virus neutralization assay

Pseudotyped vesicular stomatitis virus (VSVs) bearing SARS-CoV-2 S proteins was generated as previously described (11). The expression plasmids for the truncated S protein of SARS-CoV-2 and pCAG-SARS-CoV-2 S (Wuhan) were provided by Dr. Shuetsu Fukushi of the National Institute of Infectious Diseases, Japan. pCAGG-pm3-SARS2-Shu-d19-B1.617.2 (Delta-derived variant), pCAGG-pm3-SARS2-Shu-d19-B1.1.529.1 (Omicron BA.1-derived variant), pCAGG-pm3-SARS2-Shu-d19-B1.1.529.2 (Omicron BA.2-derived variant), and pCAGG-pm3-SARS2-Shu-d19-B1.1.529.5 (Omicron BA.5-derived variant) were also generated. VSVs bearing envelope (G) (VSV-G) were also generated. The pseudotyped VSVs were stored at − 80°C until subsequent use.

The neutralizing effects of each sample against pseudotyped viruses were examined using a high throughput chemiluminescence reduction neutralization test (htCRNT) as previously described (12). Briefly, serum was diluted 100-fold or 1600-fold with Dulbecco’s modified Eagle’s medium (DMEM; Nacalai Tesque, Inc., Kyoto, Japan) containing 10% heat-inactivated fetal bovine serum and incubated with pseudotyped SARS-CoV-2 for 1 h. After incubation, the VeroE6/TMPRSS2 cells (JCRB1819) were treated with DMEM-containing serum and pseudotyped viruses. The infectivity of the pseudotyped viruses was determined by measuring luciferase activity after 24 h of incubation at 37°C. The values of samples without pseudotyped virus and those with pseudotyped virus but without serum were defined as 0% and 100% infection (100% and 0% inhibition), respectively.

To measure the half-maximal neutralizing titer (NT_50_) values, the pooled samples were serially diluted by mixing equal volumes in a single tube, and neutralization activity was measured in duplicate by htCRNT. NT_50_ was defined as the maximum serum dilution that indicated >50% inhibition.

### SARS-CoV-2 live virus neutralization assay

SARS-CoV-2 isolates, WT (A, GISAID EPI ISL:408667), Alpha (B.1.1.7, GISAID EPI ISL:804007), Beta (B.1.351, GISAID EPI ISL:1123289), Gamma (P.1, GISAID EPI ISL:877769), Delta (B.1.617.2, GISAID EPI ISL:2158617), Omicron sub-lineage BA.1 (BA.1.1, GISAID EPI ISL:7571618), Omicron sub-lineage BA.2 (BA.2, GISAID EPI ISL:9595859), and Omicron sub-lineage BA.5 (BA.5, GISAID EPI ISL:13241867) were kindly provided by the National Institute of Infectious Diseases (Japan). To obtain a high titer of the virus stock, VeroE6/TMPRSS2 cells were infected with SARS-CoV-2 isolates and incubated in DMEM. The cell culture medium was harvested and centrifuged, and the virus-containing supernatant was stored at − 80°C after 2 or 3 days of inoculation. Prior to neutralization experiments, viral titers were verified using median tissue culture infectious dose 50 (TCID_50_) tests.

Using the same pooled samples as the pseudotyped virus assay, the serum infection-neutralization capacity was analyzed in duplicate by testing two-fold serial dilutions of sera, starting at 1/20, with 50 TCID_50_ of the virus in VeroE6/TMPRSS2 cells in four wells of 96-well plates under biosafety level 3 conditions. After 4 or 5 days of incubation, cells were fixed with paraformaldehyde and stained with an aqueous crystal violet methanol solution. The serum titer (ID_50_) that showed 50% protection from virus-induced cytopathic effects was considered to contain neutralizing antibodies and was defined as the reciprocal value of the sample dilution. Each run included an uninfected cell control, an infected cell control, and virus back-titration to confirm the virus inoculum.

### Anti-RBD and anti-nucleocapsid antibody measurements

The concentration of the anti-receptor-binding domain (RBD) antibody in serum samples was measured using the Elecsys Anti-SARS-CoV-2 S immunoassay (Roche Diagnostics GmbH, Basel, Switzerland) at Toyama University Hospital. At 2wA2D, 3mA3D, and 6mA3D, the upper limit of quantification was 25000.0 U/mL and measurements of >25000.0 U/mL were considered as 25000.0 U/mL for further statistical calculations. However, at 2wA4D, most serum samples exceeded the upper limit of quantification; therefore, such samples were diluted four-fold manually beforehand and then remeasured. Measurements of >100000.0 U/mL at 2wA4D were considered as 100000.0 U/mL for further statistical calculations. The serum concentration of anti-SARS-CoV-2 nucleocapsid antibodies was measured using an Elecsys Anti-SARS-CoV-2 immunoassay (Roche Diagnostics GmbH, Basel, Switzerland). Anti-nucleocapsid results were expressed as a cut-off index value; values ≥1.0 were considered positive for anti-nucleocapsid antibodies.

### Vaccine-related symptoms after the fourth second booster dose of vaccination

Data on adverse effects after the second booster dose (mRNA-1273 or BNT162b2 bivalent vaccine) were obtained using the same questionnaire when blood samples were collected at 2wA4D, as previously described (10). The data of participants’ age and sex were extracted from the database from the previous study (10). Items on the following adverse effects post vaccination were included: local (pain at the injection site, redness, swelling, hardness, local muscle pain, feeling of warmth, itching, and others) and systemic (fever ≥ 37.5 °C, general fatigue, headache, nasal discharge, abdominal pain, nausea, diarrhea, myalgia, joint pain, swelling of the lips and face, hives, cough, and others) symptoms.

### Statistical analysis

Statistical analysis was performed using the Mann–Whitney test to compare non-parametric groups. Friedman’s test with Dunn’s test was used for multiple comparisons among the three paired groups. Correlations between the test findings were expressed using Pearson’s correlation coefficient. Data were analyzed using GraphPad Prism version 9.4.1 (GraphPad Software, San Diego, CA). Statistical significance between different groups was defined as p < 0.05. Data are expressed as median with interquartile range (IQRs).

### Ethics approval

This study was performed in accordance with the Declaration of Helsinki and approved by the ethical review board of the University of Toyama (approval No.: R2019167). Written informed consent was obtained from all participants.

## RESULTS

### Study flow chart

Anti-RBD antibodies and neutralization activity against Omicron variant BA.1 were prevalent when the third dose of vaccination was promoted in Japan, were initially measured in 565, 425, and 321 participants at 2wA3D, 3mA3D, and 6mA3D (**Fig. 1**). To compare participants with a clarified vaccine history and known immune status, 203 participants who were assessed for humoral immunity at least one point before the second booster dose were eligible for the present study. After second booster vaccination, the participants who provided blood samples at 2wA4D were divided into two groups: the original WT vaccine group (consisted of 168 participants who received the prototype mRNA-1273 vaccine as the second booster, and the other was a bivalent Omicron BA.1-containing vaccine group (WT+BA.1 group) consisting of 23 participants who received the recently authorized Pfizer BNT162b2 bivalent (WT/Omicron BA.1) vaccine.

**Figure 1.**
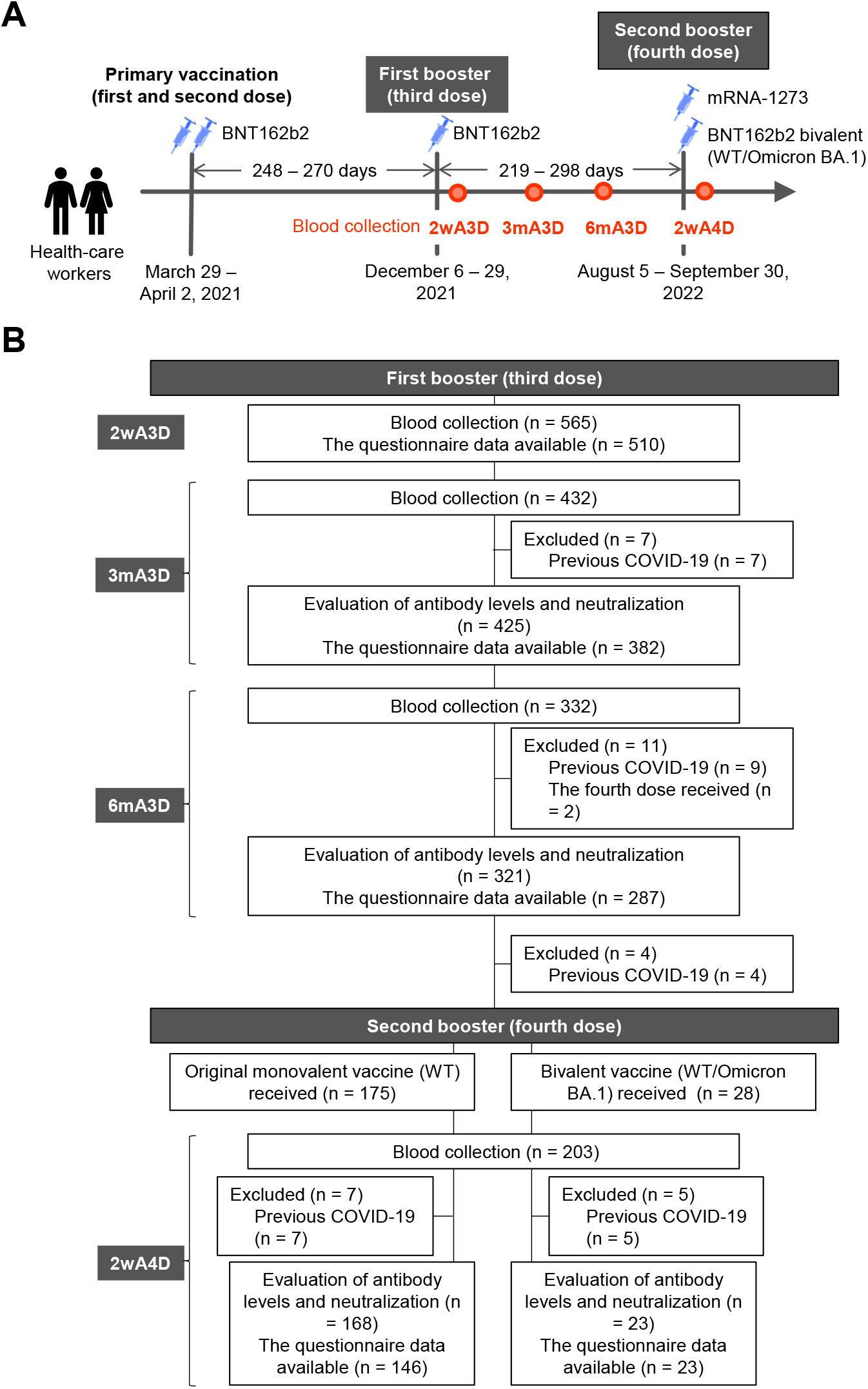
Overview of the study. **(A)** A schematic illustration of the time course of the study. **(B)** Study flow chart. Participants (n = 565) who received primary and the first booster vaccination of the BNT162b2 provided blood samples and their vaccine-induced antibody responses were assessed at 2wA3D. They were subsequently followed up at 3mA3D and 6mA3D, and those with previous SARS-CoV-2 infection were excluded. A total of 203 participants who provided blood samples and whose antibody responses were assessed at a least one point after the third dose were eligible. The participants were divided into two groups after the second booster vaccination; the Wild-type (WT) group consisting of 168 participants and the bivalent vaccine (WT+BA.1) group consisting of 23 participants. 2wA3D, 2 weeks after the third dose; 3mA3D, 3 months after the third dose; 6mA3D, 6 months after the third dose; 2wA4D, 2 weeks after the fourth dose.

### Antibody quantification and neutralizing activity before and after the second booster dose

We previously reported the immunogenicity and safety of the BNT162b2 first booster vaccine at 2wA3D in the same participants as in the present study. We assessed the durability of the first booster-acquired antibody quantification and neutralizing activity. At 3mA3D and 6mA3D, the median concentration of anti-RBD antibody was 8955.0 U/mL (IQR: 5879.0–13612.0 U/mL) and 5085.0 U/mL (IQR: 2993.0–7719.0 U/mL), and the median htCRNT value for Omicron BA.1 was 85.8% (IQR: 75.2–91.6%) and 84.1% (IQR: 64.4–91.9%) (**Fig. S1A and S1B**).

As for the second booster vaccination, there were no significant differences in the demographic and immunological characteristics and humoral immunity levels, including anti-RBD antibody and neutralization efficacy by htCRNT, between the WT and WT+BA.1 groups (**Table 1**).

**Table 1.**
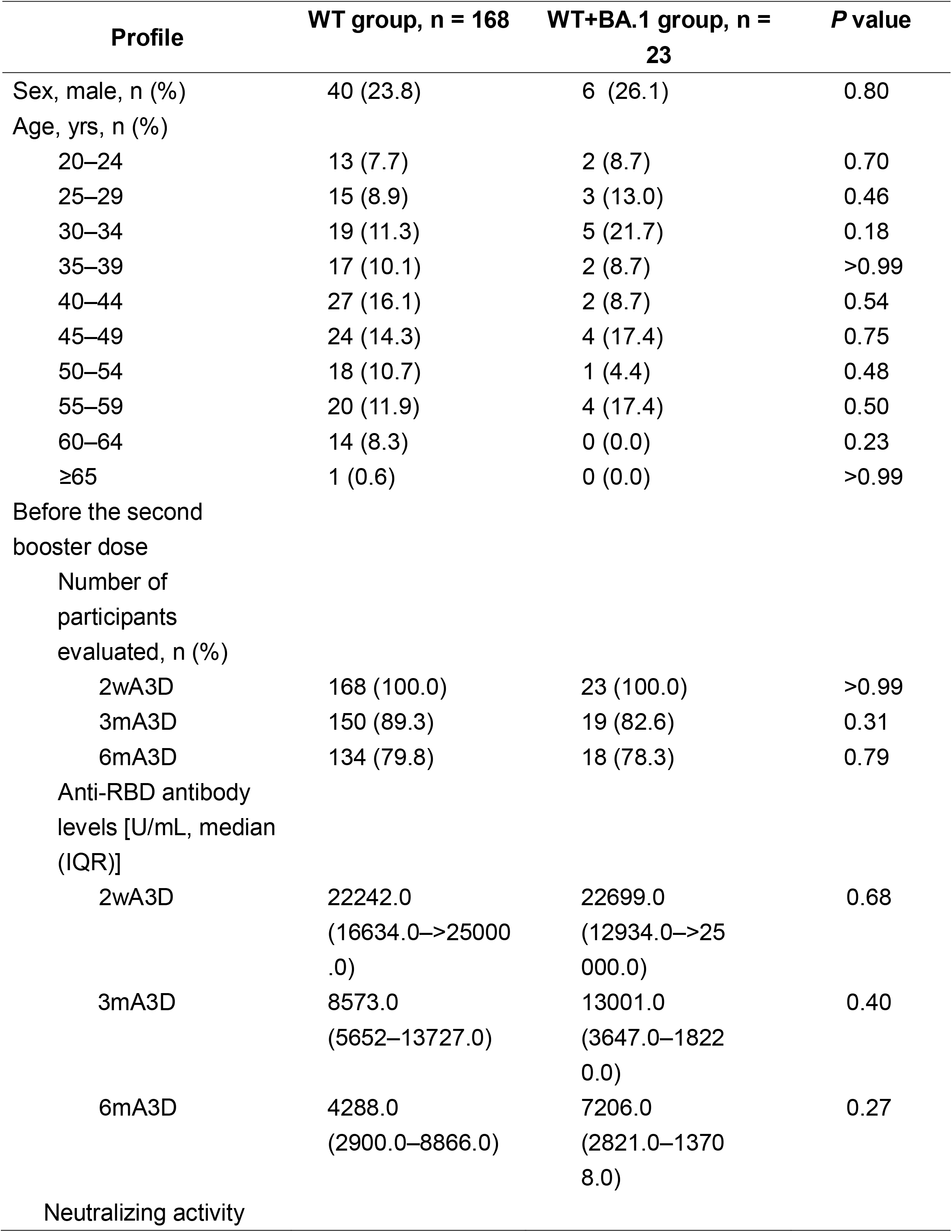
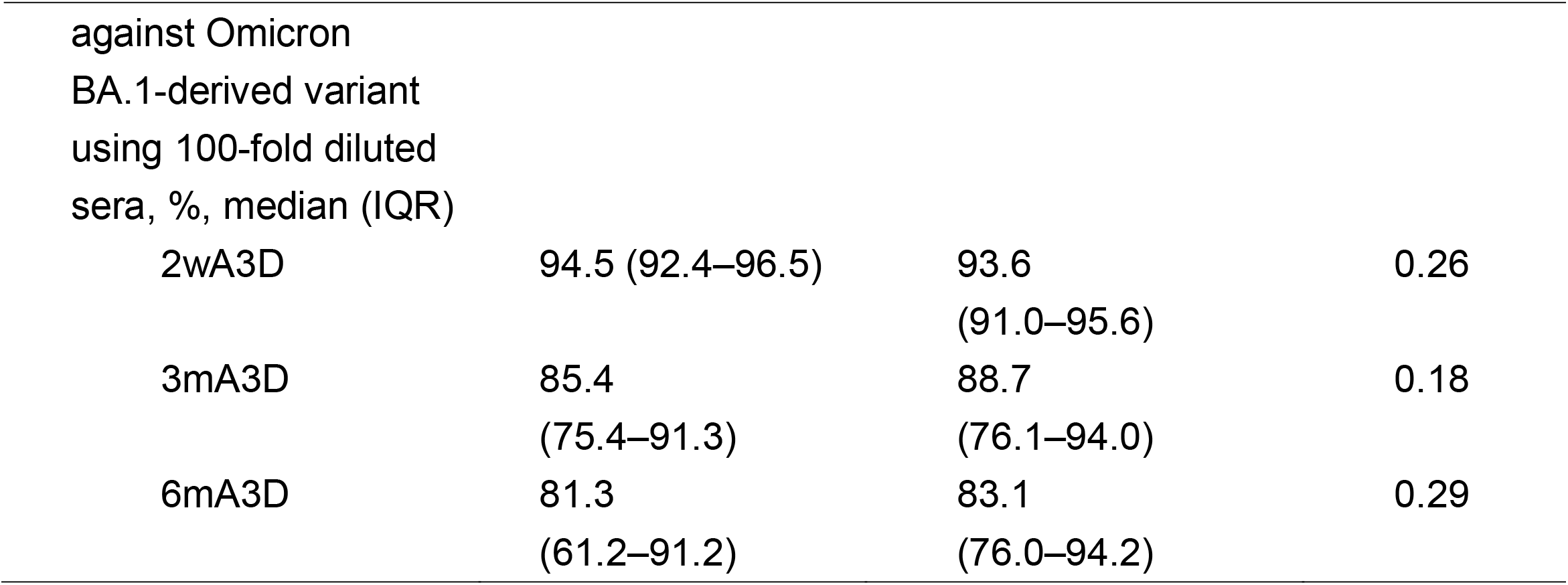
Demographic and immunological characteristics of the study participants in the original mRNA-1273 group (WT group) and BNT162b2 bivalent (WT/Omicron BA.1) vaccine group (WT+BA.1 group).

| Profile | WT group, n = 168 | WT+BA.1 group, n = 23 | P value |
| --- | --- | --- | --- |
| Sex, male, n (%) | 40 (23.8) | 6 (26.1) | 0.80 |
| Age, yrs, n (%) |  |  |  |
| 20–24 | 13 (7.7) | 2 (8.7) | 0.70 |
| 25–29 | 15 (8.9) | 3 (13.0) | 0.46 |
| 30–34 | 19 (11.3) | 5 (21.7) | 0.18 |
| 35–39 | 17 (10.1) | 2 (8.7) | >0.99 |
| 40–44 | 27 (16.1) | 2 (8.7) | 0.54 |
| 45–49 | 24 (14.3) | 4 (17.4) | 0.75 |
| 50–54 | 18 (10.7) | 1 (4.4) | 0.48 |
| 55–59 | 20 (11.9) | 4 (17.4) | 0.50 |
| 60–64 | 14 (8.3) | 0 (0.0) | 0.23 |
| ≥65 | 1 (0.6) | 0 (0.0) | >0.99 |
| Before the second booster dose |  |  |  |
| Number of participants evaluated, n (%) |  |  |  |
| 2wA3D | 168 (100.0) | 23 (100.0) | >0.99 |
| 3mA3D | 150 (89.3) | 19 (82.6) | 0.31 |
| 6mA3D | 134 (79.8) | 18 (78.3) | 0.79 |
| Anti-RBD antibody levels [U/mL, median (IQR)] |  |  |  |
| 2wA3D | 22242.0<br>(16634.0–>25000.0) | 22699.0<br>(12934.0–>25000.0) | 0.68 |
| 3mA3D | 8573.0<br>(5652–13727.0) | 13001.0<br>(3647.0–18220.0) | 0.40 |
| 6mA3D | 4288.0<br>(2900.0–8866.0) | 7206.0<br>(2821.0–13708.0) | 0.27 |
| Neutralizing activity |  |  |  |
| against Omicron<br>BA.1-derived variant<br>using 100-fold diluted<br>sera, %, median (IQR) |  |  |  |
| 2wA3D | 94.5 (92.4–96.5) | 93.6<br>(91.0–95.6) | 0.26 |
| 3mA3D | 85.4<br>(75.4–91.3) | 88.7<br>(76.1–94.0) | 0.18 |
| 6mA3D | 81.3<br>(61.2–91.2) | 83.1<br>(76.0–94.2) | 0.29 |

At 2wA4D, the median concentration of anti-RBD antibody were 26262.0 U/mL (IQR: 16951.0–38137.0 U/mL) and 24840.0 U/mL (IQR: 14828.0–41460.0 U/mL) in the WT and WT+BA.1 group, respectively (**Fig. 2A**). For neutralization against Omicron sub-variants (**Fig. 2B**), the htCRNT values using 100-fold sera against BA.2 in the WT+BA.1 group (median >99.9% [IQR: 99.9–>99.9%]) were significantly higher than those in the WT group (99.3% [IQR: 97.4–99.8%]), while there was no difference against BA.5 (WT group: 99.3% [IQR: 98.4–99.6%], vs. WT+BA.1 group; 99.5% [IQR: 97.7–99.9%]). However, significant differences were more clearly observed in the values using 1600-fold diluted sera against BA.2 (WT group: 38.5% [IQR: 10.0–61.8%] vs. WT+BA.1 group; 63.4% [IQR: 31.4–75.6%]) and BA.5 (WT group; 0.2% [IQR: 0–31.2%] vs. 22.7% [IQR: 14.3–41.0%]).

**Figure 2.**
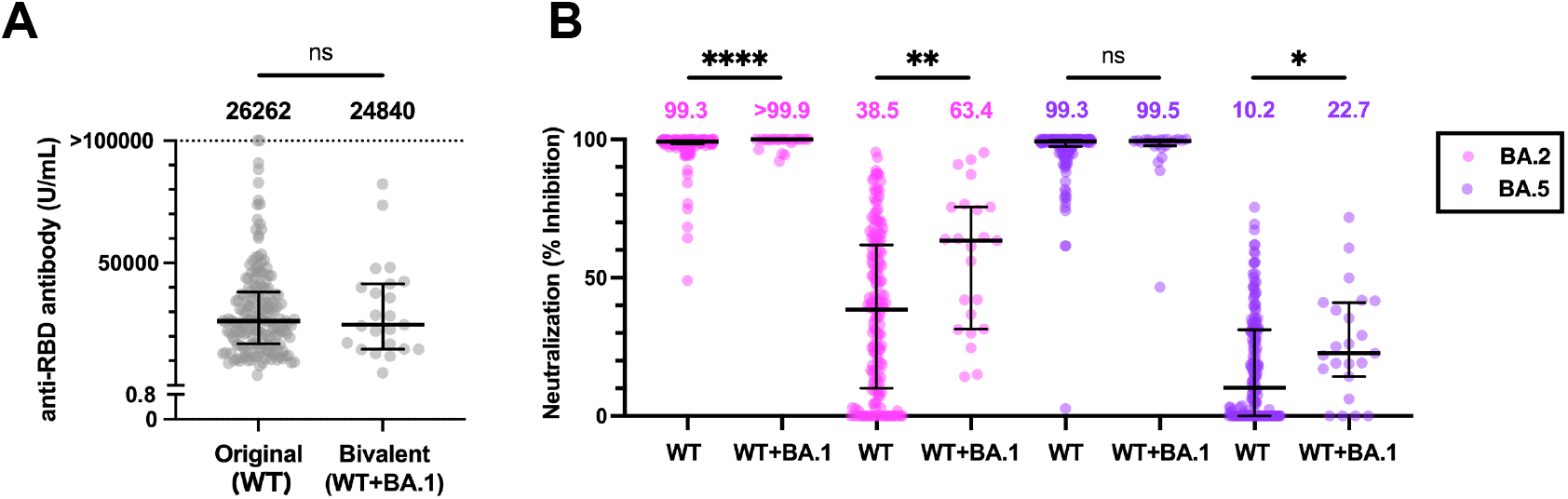
Anti-RBD antibody levels and neutralizing activity after the second booster. **(A)** Serum concentration of anti-RBD antibody at 2wA4D in the original WT group (n = 168) and the bivalent WT+BA.1 group (n = 23). Each dot represents an individual result. **(B)** Pseudotypevirus-based neutralizing activity against Omicron BA.2 and BA.5 at 2wA4D in the WT (n = 168) and the WT+BA.1 group (n = 23) The assay was performed using 100- or 1600-fold diluted serum. The numbers at the top indicate the median neutralizing values of each group. RBD, receptor-binding domain; 2wA4D, 2 weeks after the fourth dose; *, p < 0.05; **, p < 0.01; ****, p < 0.0001; ns, not significant. Bars indicate medians with interquartile ranges.

The pseudotyped virus-based NT_50_ values using pooled sera against WT, Omicron BA.1, BA.2, and BA.5 were × 400, × 400, × 100, and × 100 at 3mA3D, and × 100, × 100, × 100, and < × 100 at 6mA3D, respectively (**Fig. 3A**). At 2wA4D, neutralization was restored against all evaluated pseudotyped viruses in the WT and WT+BA.1 groups, and the NT_50_ values for WT, Omicron BA.1, BA.2, and BA.5 were 1600, 1600, 1600, and 400, respectively. The WT+BA.1 group at 2wA4D showed higher NT_50_ values against BA.5 than did the WT group.

**Figure 3.**
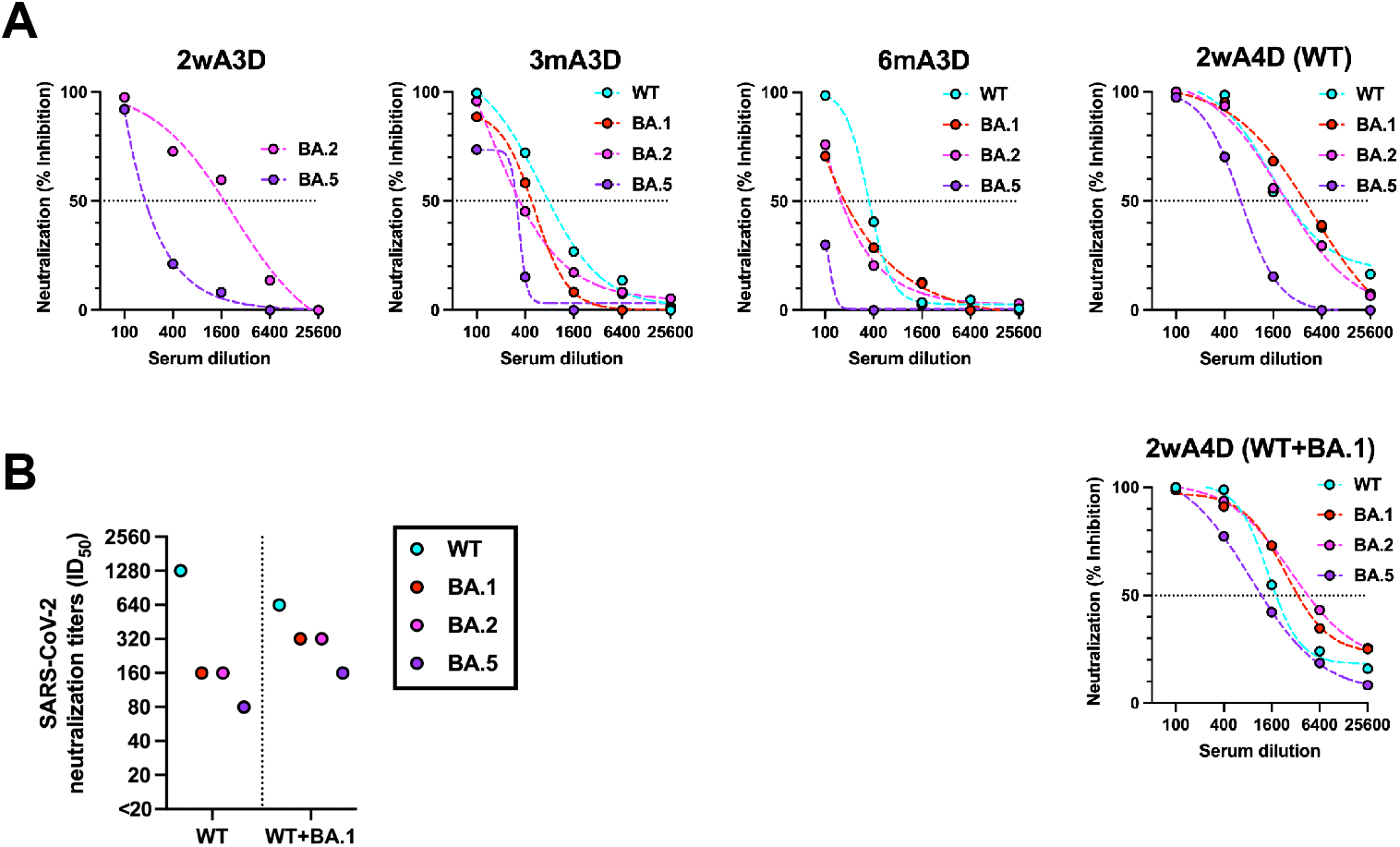
Neutralizing activities before and after the second booster dose. **(A)** Neutralization titers (NT_50_) against WT, Omicron BA.1, BA.2, and BA.5-pseudotyped viruses at 2wA3D (n = 565), 3mA3D (n = 425), 6mA3D (n = 321), and 2wA4D in the WT (n = 168) and the WT+BA.1 group (n = 23) using the pooled serum. Dotted lines indicate interpolate standard curves. **(B)** ID_50_ titers against live viruses at 2wA4D in the WT (n = 168) and in the WT+BA.1 group (n = 23). The assay was performed by using pooled serum. WT, wild-type; 2wA3D, 2 weeks after the third dose; 3mA3D, 3 months after the third dose; 6mA3D, 6 months after the third dose; 2wA4D, 2 weeks after the fourth dose.

Live virus-based neutralization at 2wA4D also showed higher ID_50_ titers against Omicron sub-lineage BA.1, BA.2, and BA.5 in the WT+BA.1 group than in the WT group, while ID_50_ against WT in the WT+BA.1 group was lower than that in the WT group (**Fig. 3B**). The second booster recovered the neutralization activity, which gradually decreased after the first booster (2wA3D, 3mA3D, and 3mA3D), and the WT+BA.1 group showed higher ID_50_ values against Omicron sub-lineages than those at 2wA3D (**Fig. S2**).

### Vaccine-related symptoms after the first and second booster

Vaccine-related symptoms after the second booster dose were collected from participants in the WT and the WT+BA.1 groups (**Table 2**). Local and systemic symptoms were similarly observed in the WT and WT+BA.1 groups, with percentages of 94.5% and 84.3% in the WT group and 100% and 82.6% in the WT+BA.1 group, respectively. Frequent local reactions included pain at the injection site and local muscle pain; the most frequent systemic reactions were general fatigue, fever ≥ 37.5 °C, headache, and joint pain in both groups. Among the symptoms collected in the questionnaire, the incidence of diarrhea was significantly higher in the WT+BA.1 (13.0%) group than in the WT group (2.1%).

**Table 2.** Vaccine-related symptoms after the fourth dose of vaccination.

| **Profile** | **WT group, n = 146** | **WT+BA.1 vaccine group, n = 23** | **P value** |
| --- | --- | --- | --- |
| Symptom, n (%) |  |  |  |
| Local symptom | 138 (94.5) | 23 (100.0) | 0.60 |
| Pain at injection site | 109 (74.7) | 21 (91.3) | 0.11 |
| Redness | 25 (17.1) | 2 (8.7) | 0.54 |
| Swelling | 27 (18.5) | 1 (4.4) | 0.13 |
| Hardness | 11 (7.5) | 1 (4.4) | >0.99 |
| Local muscle pain | 68 (46.6) | 13 (56.5) | 0.50 |
| Feeling of warmth | 33 (22.6) | 3 (13.0) | 0.41 |
| Itching | 12 (8.2) | 0 (0.0) | 0.37 |
| Others | 3 (2.1) | 1 (4.4) | 0.45 |
| Systemic symptom | 123 (84.3) | 19 (82.6) | 0.77 |
| Fever $\geq 37.5$ °C | 61 (41.8) | 9 (39.1) | >0.99 |
| General fatigue | 109 (74.7) | 16 (70.0) | 0.61 |
| Headache | 58 (39.7) | 7 (30.4) | 0.49 |
| Nasal discharge | 3 (2.1) | 1 (4.4) | 0.45 |
| Abdominal pain | 2 (1.4) | 2 (8.7) | 0.090 |
| Nausea | 13 (8.9) | 2 (8.7) | >0.99 |
| Diarrhea | 3 (2.1) | 3 (13.0) | 0.034 |
| Myalgia | 21 (14.4) | 1 (4.4) | 0.32 |
| Joint pain | 37 (25.3) | 5 (21.7) | 0.80 |
| Swelling of the lips and face | 0 (0.0) | 0 (0.0) | >0.99 |
| Hives | 0 (0.0) | 0 (0.0) | >0.99 |
| Cough | 1 (0.7) | 0 (0.0) | >0.99 |
| Others | 9 (6.2) | 3 (12.7) | 0.21 |

Since immunogenicity after the first booster dose was maintained at a higher level in those who exhibited symptoms (**Fig. S4A and S4B**), the relationship between vaccine-related symptoms and humoral immunity after the second booster dose was investigated. At 2wA4D in the WT group, anti-RBD antibody levels were remarkably elevated in participants who presented with fever ≥ 37.5 °C, general fatigue, and at least one systemic or local symptom (**Fig. 4A**). The neutralization against BA.2 and BA.5 was highly elevated in participants who had systemic symptoms (assay using 1600-fold sera for BA.2 and assay using 100-fold sera for BA.5), and fever ≥ 37.5 °C and general fatigue showed significant elevation in the neutralizing assay (**Fig. 4B**). In the WT+BA.1 group, an association between the symptoms and humoral immunity was not observed.

**Figure 4.**
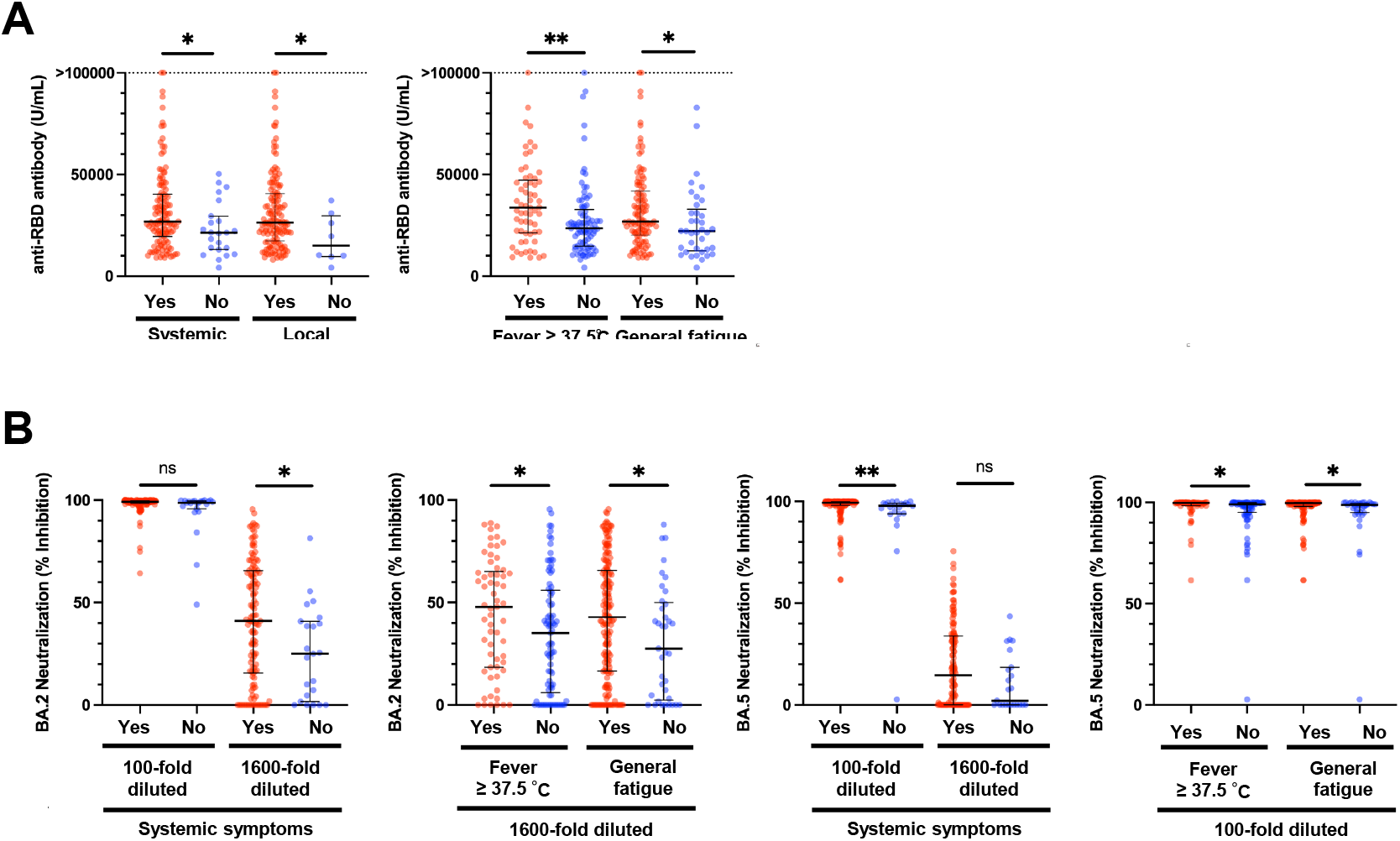
Relationship of vaccine-induced antibody levels and vaccine-related symptoms after the second booster dose in questionnaire-answered population. **(A)** Anti-RBD antibody levels in individuals with or without systemic or local symptoms (left) and specific symptoms (right) at 2wA4D in the WT group (n□=□146). **(B)** Individual neutralizing activity against BA.2 (left panels) and BA.5 (right panels). RBD, receptor-binding domain; 2wA4D, 2 weeks after the fourth dose. *, p < 0.05; **, p < 0.01; ns, not significant. Bars indicate medians with interquartile ranges.

The relationship between vaccine-related symptoms after the first and second booster doses was evaluated (**Table S1**). More than two-thirds of those who had experienced symptoms such as fever ≥ 37.5 °C, general fatigue, or at least one systemic or local symptom after the first booster dose also displayed the same symptoms after the second booster dose. In contrast, symptoms that were absent after the first booster dose were observed in 27.6–71.4% in the WT group and 10.0–50.0% in the WT+BA.1 group.

## DISCUSSION

Although Omicron BA.1-containing bivalent vaccines have been authorized, real-world data validating their safety and antibody responses remain scarce. The C4591031 substudies D and E evaluated the safety, immunogenicity, and reactogenicity of the BNT162b2 bivalent (WT/Omicron BA.1) vaccine compared to the original BNT162b2 vaccine as a second booster dose in participants aged over 55 years without previous SARS-CoV-2 infection (13). This clinical study showed the superior effectiveness of the second booster dose with the Omicron BA.1 bivalent vaccine over the original monovalent vaccine.

However, data are still insufficient ensuring safety and effectiveness of the bivalent Omicron-containing vaccines; it is thus difficult for clinicians and policy makers to decide what to choose or recommend as the booster, while considering delay of vaccination or vaccine shortages. Here, we report the first prospective longitudinal study to assess the safety, immunogenicity, and reactogenicity of a new BNT162b2 bivalent (WT/Omicron BA.1) vaccine in healthcare workers with no history of SARS-CoV-2 infection.

In the present study, the bivalent (WT+BA.1) vaccine elicited higher neutralization against Omicron BA.2 and BA.5 subvariants than the monovalent vaccine, similar to previous studies (8). BA.5 evades other WT and Omicron BA.1 antibodies (14-16). Although immunity induced by BA.1 is relatively less protective against BA.5, it provides some protection against reinfection with BA.2 and BA.5 (17, 18). Our data showed that the bivalent (WT+ BA.1) vaccine produced 1.5–2.0-fold higher NT_50_ and ID_50_ values against BA.5 than monovalent vaccines, but although still lower than those directed against other SARS-CoV-2 variants. This suggests that BA.5-neutralizing antibodies are induced by a single Omicron booster, although probably not sufficiently; the dosage therefore requires repeated boosters.

Long-term maintenance of vaccine-induced antibodies is important not only for individual protection against infection, but also for social vaccination strategies. The comparison between the results of our previous report and our current study after the first booster dose led to an inference that the original monovalent vaccine rapidly loses immunity against Omicron (10). This inference conforms with reports based on similar studies (14, 19). In fact, vaccine effectiveness waned remarkably within a few months of administration in clinical settings (20, 21). Therefore, even when the second booster dose shows temporary recovery, it is expected to decline in a similar manner. In contrast, for the bivalent (WT/BA.1) vaccine, an ongoing prospective longitudinal study should be performed because it is the only way to determine the persistence of humoral immunity levels after the second booster dose.

The evaluation of vaccine-related reactions is important for the continuation of repeated vaccinations. The frequency of adverse events after the second booster dose was similar to those after the first booster dose (8, 10). Similar to our previous studies about the vaccine efficacy (10, 22), a positive relationship was observed between higher antibody responses and some specific adverse reactions. Furthermore, many participants who experienced symptoms after the first booster dose showed the same symptoms after the second booster dose. In contrast, those who did showed symptoms after the first booster dose were less likely to experience the same adverse symptoms at the second booster dose. These findings suggest that acquiring immunity from vaccination is somewhat related to vaccine-related adverse events, and that there are individual differences.

This study’s chief limitation is that the targeted ages were limited to 20–69 years old and simplified background information was collected. Thus, the antibody responses in older and younger individuals are unknown. In addition, since detailed interviews regarding specific underlying diseases and medications were not conducted, it was impossible to infer the efficacy modified by host health. Second, the number of participants in the BNT162b2 bivalent vaccine group was small because most HCWs had already received the second booster dose of vaccine through vaccine campaigns before the bivalent vaccine was authorized.

In conclusion, the second booster vaccination of the bivalent Omicron BA.1-containing BNT162b2 vaccine induced higher neutralizing activity against BA.2 and BA.5 than those of the prototype mRNA-1273 vaccine.

## Data Availability

The authors confirm that data supporting the findings of this study are available within the article.

## Conflicts of interest

The authors have no conflicts of interest to declare.

## Funding

This study was supported by the Research Program on Emerging and Re-emerging Infectious Diseases from the AMED (grant number JP20he0622035) (YM, HT, and YY) and (grant number JP21fk0108588) (YM and HT), a research funding grant from the president of the University of Toyama (YM, NH, and YY), Toyama Pharmaceutical Valley Development Consortium (YM, NH, and YY), Morinomiyako Medical Research Foundation (HK), Kurozumi Medical Foundation (HK), The Hokuriku Bank grant-in-aid for Young Scientists (HK), and Japan Society for the Promotion of Science (JSPS) KAKENHI grant number JP22K20768 (HK). The funding bodies played no role in the design of the study, collection, analysis, or interpretation of data, nor in writing the manuscript.

## Acknowledgments

We thank the staff at Toyama University Hospital for their help in collecting the specimens and questionnaires.

---

**Table S1.**
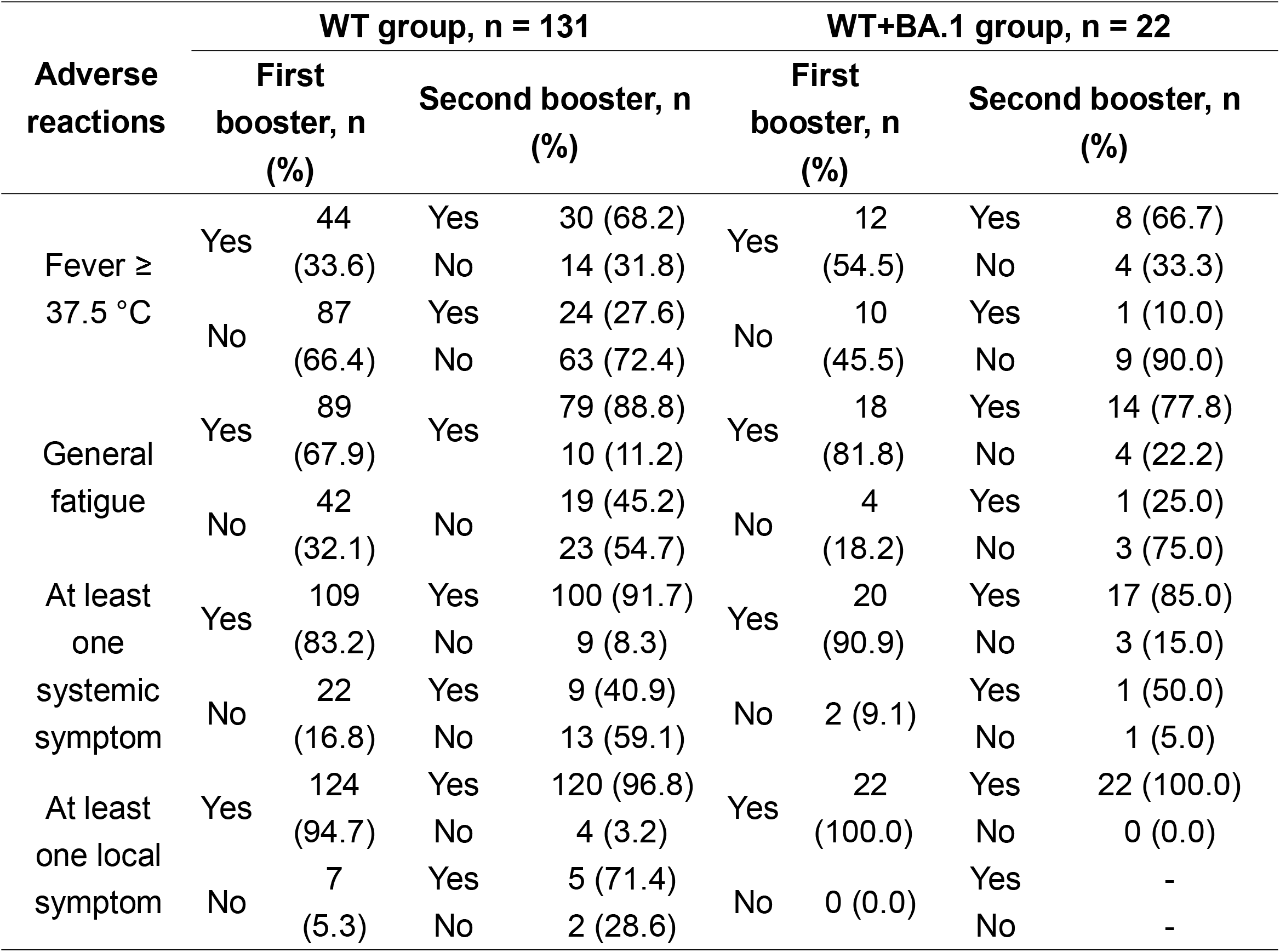
Relationship between vaccine-related symptoms after the first and second booster.

## Figure legends

**Supplemental Figure 1 Anti-RBD antibody levels and neutralizing activity before the second booster dose**

**(A)** Serum concentration of anti-RBD antibody at 3mA3D (n = 425) and 6mA3D (n = 321). Each dot represents an individual result. **(B)** Individual neutralizing activity against Omicron BA.1-pseudotyped virus at 3mA3D (n = 425) and 6mA3D (n = 321) using 100-fold diluted serum. The numbers at the top indicate the median neutralizing value for each group.

RBD, receptor-binding domain; 3mA3D, 3 months after the third dose; 6mA3D, 6 months after the third dose; *, p < 0.05; ****, p < 0.0001. Bars indicate medians with interquartile ranges.

**Supplemental Figure 2 ID_50_ titers against live Wild-type-, Omicron BA.1, BA.2, and BA.5 viruses at 2wA3D, 3mA3D, and 6mA3D**

ID_50_ titers against live SARS-CoV-2 viruses, WT, Omicron BA.2, and BA.5 at 2wA3D (n = 565), 3mA3D (n = 425), and 6mA3D (n = 321) using the pooled serum.

2wA3D, 2 weeks after the third dose; 3mA3D, 3 months after the third dose; 6mA3D, 6 months after the third dose; 2wA4D, 2 weeks after the fourth dose.

**Supplemental Figure 3 Relationship of vaccine-induced antibody levels and vaccine-related symptoms before the second booster dose in questionnaire-answered population**

**(A)** Relationship between anti-RBD antibody levels and vaccine-related responses including fever ≥ 37.5 °C, general fatigue, systemic, and local symptoms at 3mA3D (n□=□425) and 6mA3D (n = 321). **(B)** Relationship between neutralizing activity against the Omicron BA.1-pseudotyped virus and vaccine-related symptoms.

RBD, receptor-binding domain; 3mA3D, 3 months after the third dose; 6mA3D, 6 months after the third dose; *, p < 0.05; **, p < 0.01; ***, p < 0.001; ****, p < 0.0001. Each dot represents an individual result. Bars indicate medians with interquartile ranges.

## Notes

### Competing Interest Statement

The authors have declared no competing interest.

### Author Declarations

This study was performed in accordance with the Declaration of Helsinki and approved by the ethical review board of the University of Toyama (approval No.: R2019167).

