## Supplementary figures and images for "Efficacy of the wild-type/Omicron BA.1 bivalent vaccine as the second booster dose against Omicron BA.2 and BA.5"

### Fig. S1

#### Slide 1
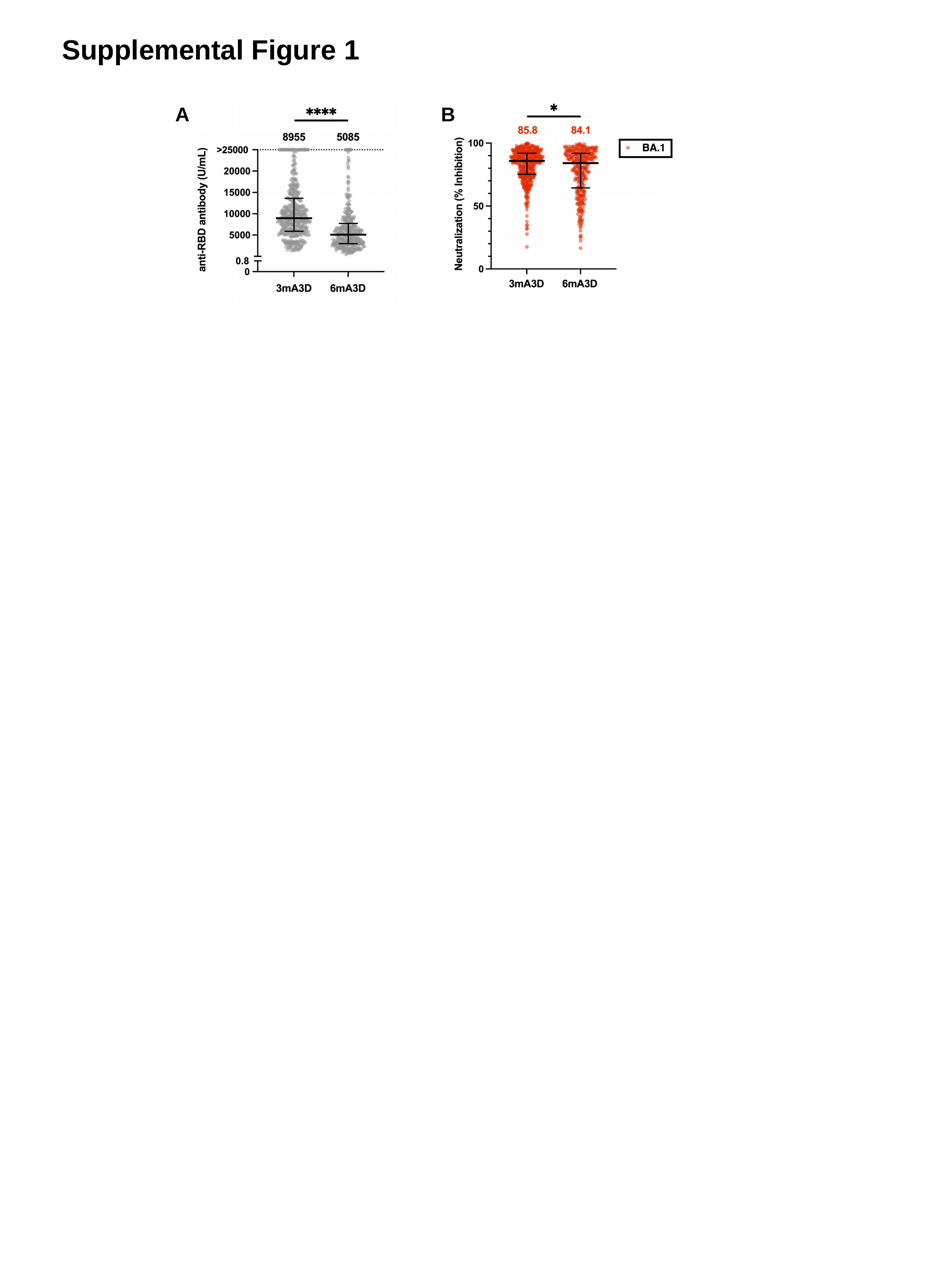

Supplemental Figure 1
A
B

### Fig. S2

#### Slide 1
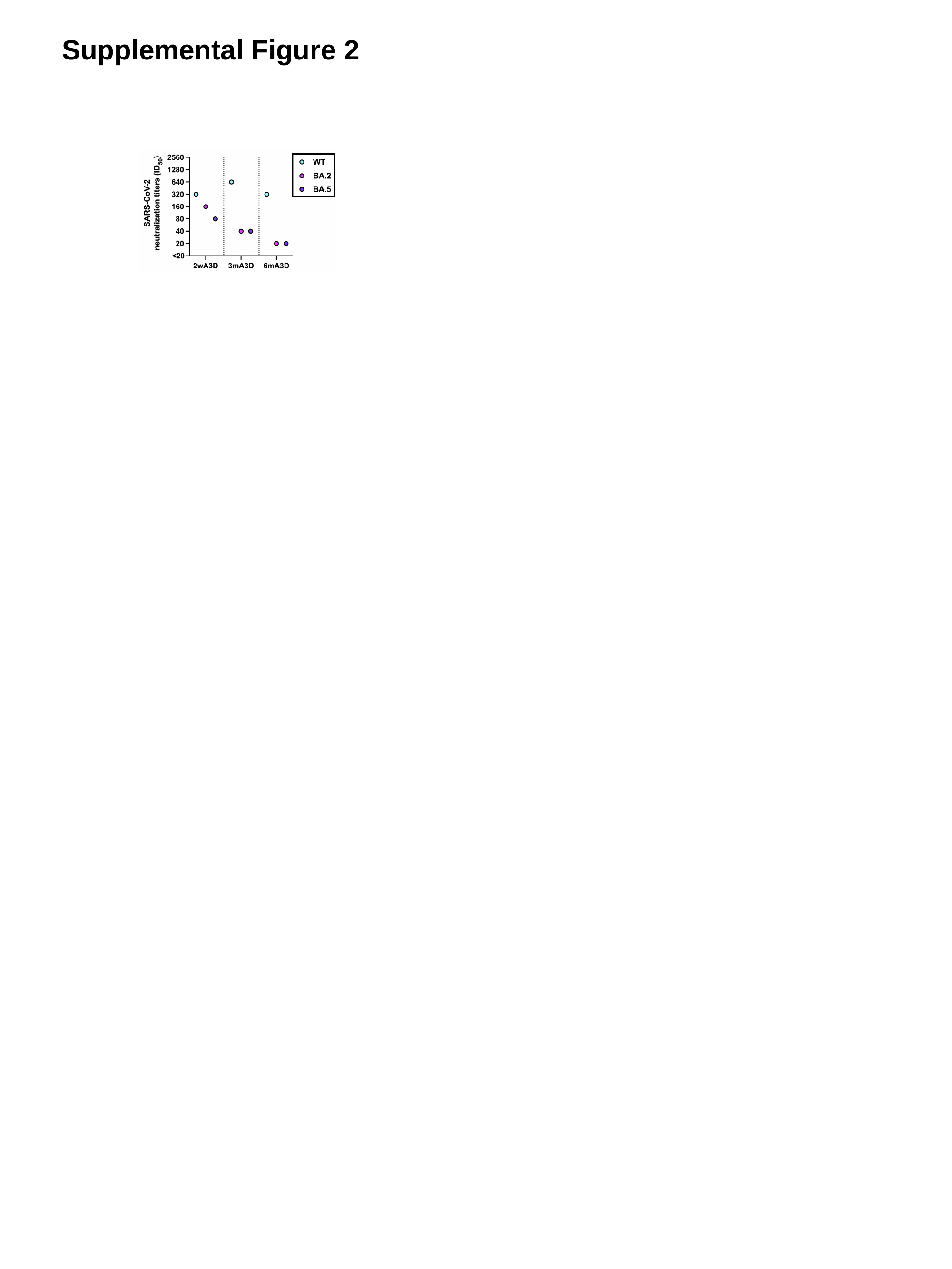

Supplemental Figure 2
