## Supplementary material for "Efficacy of the wild-type/Omicron BA.1 bivalent vaccine as the second booster dose against Omicron BA.2 and BA.5": Fig. S3

### Slide 1
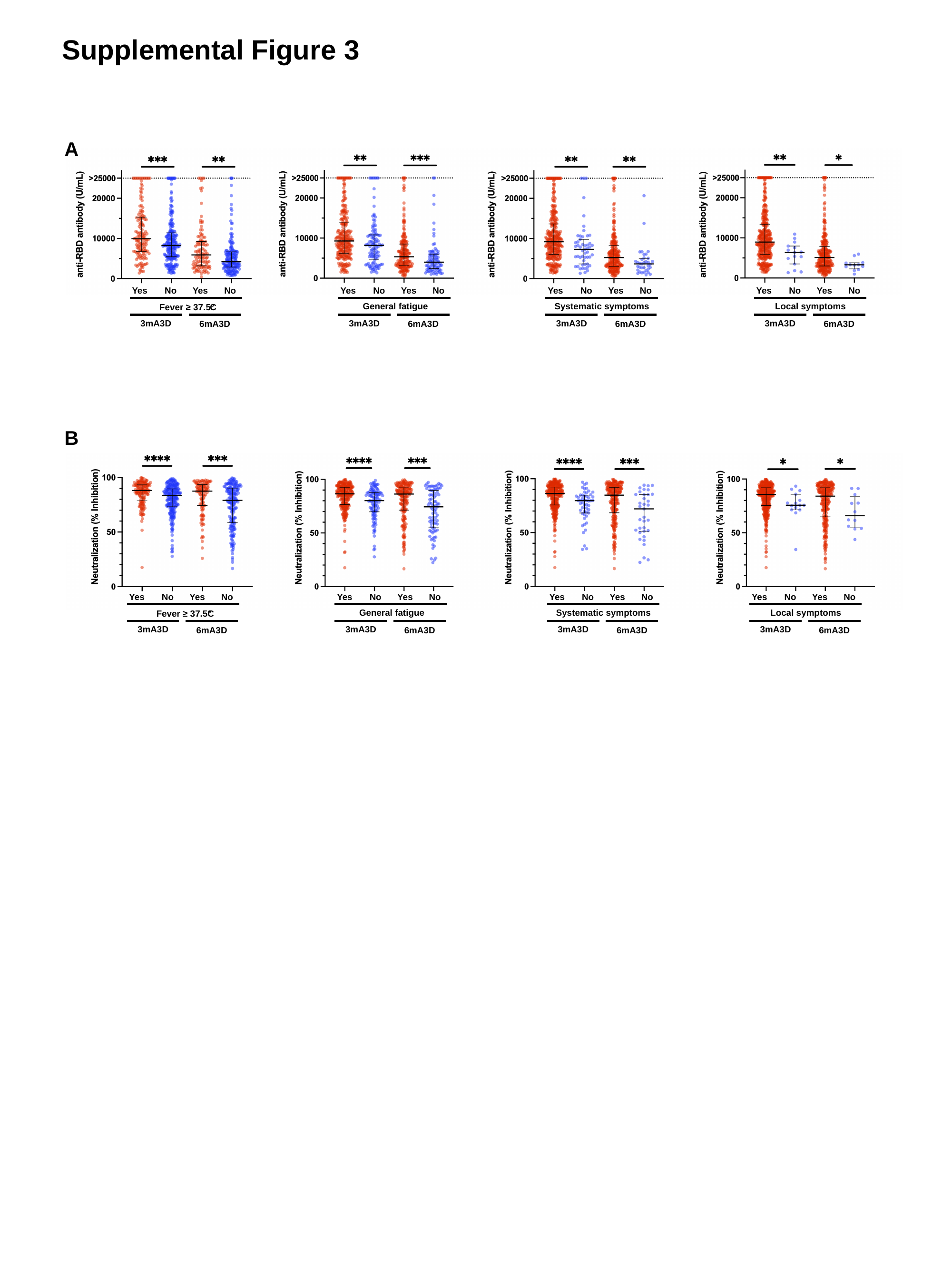

Supplemental Figure 3
A
No
No
Yes
Yes
Fever ≥ 37.5°
C
3mA3D
6mA3D
No
No
Yes
Yes
General fatigue
3mA3D
6mA3D
No
No
Yes
Yes
Systematic symptoms
3mA3D
6mA3D
No
No
Yes
Yes
Local symptoms
3mA3D
6mA3D
B
No
No
Yes
Yes
Systematic symptoms
3mA3D
6mA3D
No
No
Yes
Yes
Fever ≥ 37.5°
C
3mA3D
6mA3D
No
No
Yes
Yes
General fatigue
3mA3D
6mA3D
No
No
Yes
Yes
Local symptoms
3mA3D
6mA3D
